# Blood protein levels predict leading incident diseases and mortality in UK Biobank

**DOI:** 10.1101/2023.05.01.23288879

**Authors:** Danni A. Gadd, Robert F. Hillary, Zhana Kuncheva, Tasos Mangelis, Yipeng Cheng, Manju Dissanayake, Romi Admanit, Jake Gagnon, Tinchi Lin, Kyle Ferber, Heiko Runz, Biogen Biobank Team, Riccardo E. Marioni, Christopher N. Foley, Benjamin B. Sun

## Abstract

The circulating proteome offers insights into the biological pathways that underlie disease. Here, we test relationships between 1,468 Olink protein levels and the incidence of 23 age-related diseases and mortality, over 16 years of electronic health linkage in the UK Biobank (N=47,600). We report 3,201 associations between 961 protein levels and 21 incident outcomes, identifying proteomic indicators of multiple morbidities. Next, protein-based scores (ProteinScores) are developed using penalised Cox regression. When applied to test sets, six ProteinScores improve Area Under the Curve (AUC) estimates for the 10-year onset of incident outcomes beyond age, sex and a comprehensive set of 24 lifestyle factors, clinically-relevant biomarkers and physical measures. Furthermore, the ProteinScore for type 2 diabetes outperformed a polygenic risk score, a metabolomic score and HbA1c – a clinical marker used to monitor and diagnose type 2 diabetes. These data characterise early proteomic contributions to major age-related disease and demonstrate the value of the plasma proteome for risk stratification.

## Introduction

Omics signatures are increasingly used to hone clinical trial design ^1^, while also opening up avenues for more personalised healthcare ^2,3^. Of all the omics layers that can be measured from a single blood test, proteomics arguably holds the most intrinsic predictive potential, given that proteins are the intermediary effectors of health maintenance and disease and are often the targets of pharmacological interventions. Several studies have shown that circulating proteins can discriminate disease cases from controls and delineate risk of incident diagnoses ^4–11^. Screening the proteome against incident outcomes has been shown to identify sets of individual protein markers – some of which have then been causally-implicated in disease ^8,12–14^. This demonstrates the value protein data have in informing therapeutic targeting and reflecting the internal processes occurring in the body that precede formal diagnoses.

While singular protein markers offer insight into the mediators of disease, harnessing multiple proteins simultaneously can be expected to generate predictive tools with even greater clinical utility ^15^. Although cross-sectional case-control studies can inform on the molecular signatures of diagnosed diseases, longitudinal approaches that assess early biomarker signatures relating to time-to-disease are more suited to risk stratification. Clinically-available risk profiling scores that rely on lifestyle and health information such as QRISK and ASSIGN typically profile 10-year onset risk of disease ^16,17^. Scores such as these stratify where individuals lie on the disease-risk continuum for a population, but do not include omics features. While proteomic and metabolomics scores have been developed for certain time-to-event outcomes in isolation ^9,18–22^, these predictors are rarely developed and tested at scale. Proteomic predictors have been trained using the SomaScan platform for diabetes and cardiovascular event risk and multiple lifestyle and health indicators ^23^. Metabolomics data have been recently shown to facilitate incident disease prediction in the UK Biobank ^24^. However, no study has systematically assessed proteomic score generation for multiple incident morbidities.

Here, we quantify how large-scale proteomic sampling can identify candidate protein targets and facilitate the prediction of incident outcomes in the UK Biobank **(Fig. 1)**. We use 1,468 Olink plasma protein measurements in 47,600 individuals available as part of the UK Biobank Pharma Proteomics Project (UKB-PPP) ^25^. First, Cox proportional hazards (PH) models are used to characterise associations between each protein and 23 incident diseases, ascertained via data linkage to primary and secondary care records and mortality over 16 years of follow-up. Next, the dataset is randomly split into training and testing subsets to train proteomic scores (ProteinScores) and assess their utility for modelling either 5-year or 10-year onset of the 19 incident outcomes that had a minimum of 150 cases available. Type 2 diabetes is taken forward to explore the potential value that ProteinScores may offer, beyond clinical biomarkers, polygenic risk scores (PRS) and metabolomics measures.

**Figure 1.**
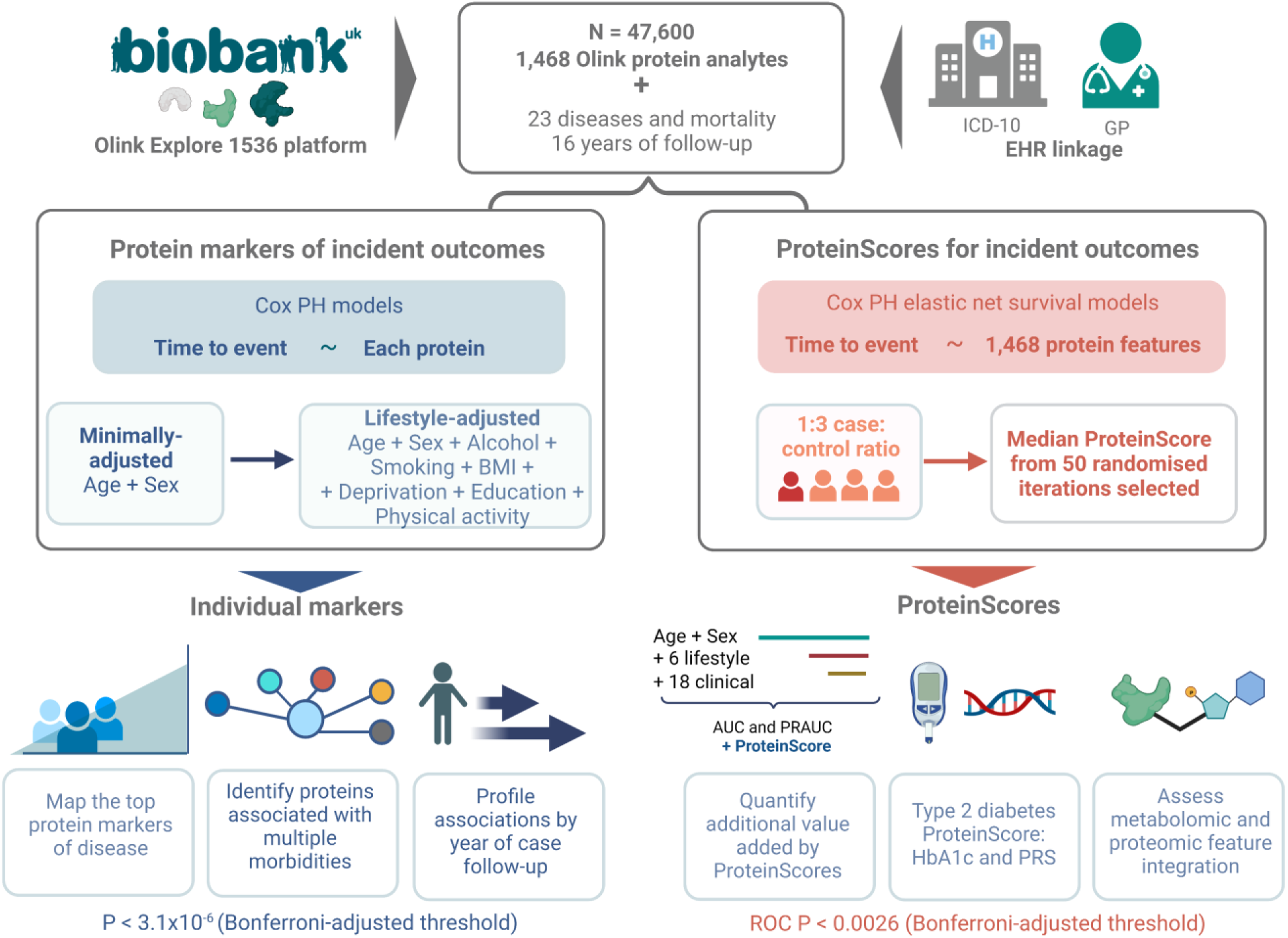
Proteomic assessment of 23 incident diseases and mortality in the UK Biobank (N=47,600). First, individual Cox proportional hazards (PH) models were used to profile relationships between baseline protein analytes and incident diseases or death, over a maximum of 16 years of electronic health linkage. Associations that had P < 3.1×10^−6^ (Bonferroni-adjusted threshold) in minimally-adjusted (age and sex) and lifestyle-adjusted models were retained. Proteins associated with multiple morbidities were identified and associations were explored by year of case follow-up. Next, proteomic predictors (ProteinScores) were trained using Cox PH elastic net regression for 19 of the incident outcomes with a minimum of 150 cases. All ProteinScores were developed for 10-year onset of disease, except endometriosis, cystitis and amyotrophic lateral sclerosis that had case distributions that were better-suited to 5-year assessment (80% of cases diagnosed by year 8 of follow-up). Of fifty ProteinScore iterations with randomly sampled train and test populations, the ProteinScore with median improvement in AUC beyond a minimally-adjusted model was selected. Improvements in AUC and PRAUC due to adding the ProteinScores into models with increasingly complex covariate structures were quantified. The type 2 diabetes trait was taken forward as a case study to explore the potential value ProteinScores may offer, in the context of HbA1c (a clinically used biomarker), a polygenic risk score (PRS) and integration of metabolomics features for scoring.

## Results

### The UKB-PPP sample

Of the 1,472 protein levels available in the UKB-PPP sample, 1,463 are unique, due to CXCL8, IL6 and TNF having multiple analyte measurements (annotation information provided in **Supplementary Table 1**). After quality control and removal of outliers, measurements for 52,744 individuals were available. In this study, a total sample of 47,600 individuals with 1,468 protein analytes was used, after exclusions for related individuals and missing data **(Supplementary Fig. 1, Methods)**. The 1,468 analyte measurements correspond to 1,459 unique protein levels. Demographic and phenotypic information is presented in **Supplementary Table 2**. Principal components analyses indicated that the first 678 components explained a cumulative variance of 90% in the protein levels **(Supplementary Table 3)**.

### Protein associations with incident outcomes

First, differential plasma protein levels that were associated with the onset of 23 diseases (that included leading causes of disability, morbidity and reductions in healthy life expectancy) ^26–28^ were identified, up to 16 years prior to formal diagnoses. Time-to-mortality was also considered as an outcome (4,446 individuals had died during the 16-year follow-up period). A total of 35,232 associations were tested (1,468 analytes and 24 outcomes). The number of cases and controls available in Cox PH models, with mean time-to-onset for cases is presented for each outcome in **Table 1**.

**Table 1.** The 24 incident outcomes profiled over a maximum of 16 years of follow-up in the UK Biobank (N=47,600). Counts for incident cases and controls are provided, with mean years to diagnosis for incident cases. These data were used in individual Cox PH models to identify protein levels that were associated with incident outcomes. ^a^ Sex-stratified traits. ^b^ Alzheimer’s and vascular dementia were restricted to individuals aged 65 years or above at the time of diagnosis for cases, or at the time or censoring for controls. CNS: central nervous system.

| Incident diagnosis | Incident cases (N) | Controls (N) | Mean years to incident case diagnosis (sd) |
| --- | --- | --- | --- |
| Schizophrenia | 54 | 47449 | 6.5 (3.4) |
| Brain/CNS cancer | 82 | 47507 | 5.5 (2.8) |
| Multiple sclerosis | 96 | 47165 | 5.6 (3.2) |
| Major depression | 111 | 47229 | 4.2 (3.1) |
| Systemic lupus erythematosus | 134 | 47096 | 5.1 (2.6) |
| Endometriosis <sup>a</sup> | 157 | 24768 | 4.8 (3.3) |
| Vascular dementia <sup>b</sup> | 195 | 33907 | 8.1 (3) |
| Gynaecological cancer <sup>a</sup> | 256 | 25185 | 5 (3) |
| Amyotrophic lateral sclerosis | 264 | 47269 | 5.4 (2.7) |
| Inflammatory bowel disease | 275 | 46727 | 5.9 (3.3) |
| Lung cancer | 403 | 47158 | 5.9 (3.2) |
| Liver disease | 432 | 47104 | 7 (3.3) |
| Alzheimer's dementia <sup>b</sup> | 446 | 33642 | 7.8 (2.8) |
| Colorectal cancer | 508 | 46890 | 5.8 (3.1) |
| Cystitis <sup>a</sup> | 531 | 24160 | 4.1 (3) |
| Rheumatoid arthritis | 593 | 46310 | 6.8 (3.2) |
| Parkinson's disease | 659 | 46802 | 5.4 (3.2) |
| Ischaemic stroke | 765 | 46657 | 6.8 (3.4) |
| Breast cancer <sup>a</sup> | 772 | 24086 | 5.2 (3.1) |
| Prostate cancer <sup>a</sup> | 1001 | 20628 | 5.7 (3.1) |
| Chronic obstructive pulmonary disease | 1998 | 44948 | 6.3 (3.4) |
| Type 2 diabetes | 2822 | 43370 | 6 (3.3) |
| Ischaemic heart disease | 3338 | 41341 | 6.3 (3.4) |
| Death | 4445 | 43155 | 7.9 (3.5) |

In minimally-adjusted (age- or age- and sex-adjusted) models, there were 5,252 associations between 1,209 unique protein analytes and 23 outcomes (Bonferroni-adjusted P threshold = 3.1×10^−6^) **(Supplementary Table 4).** Further adjustment for health and lifestyle risk factors (body mass index (BMI), alcohol consumption, social deprivation, education status, smoking status and physical activity) led to the attenuation of 2,051 of the minimally-adjusted associations, with 3,201 that remained (Bonferroni-adjusted P threshold = 3.1×10^−6^) **(Fig. 2a, Supplementary Table 5)**. The 3,201 associations involved 961 unique protein analytes and 21 outcomes, ranging from one association for amyotrophic lateral sclerosis, cystitis and multiple sclerosis, to 646 and 664 for mortality and liver disease, respectively. No associations were found for brain/CNS cancer, major depression and schizophrenia. **Supplementary Table 6** summarises the 961 unique protein analytes selected across the 3,201 associations by disease and by direction of effect (i.e. 303 associations with Hazard Ratio (HR) < 1 and 2,898 associations with HR > 1).

**Figure 2.**
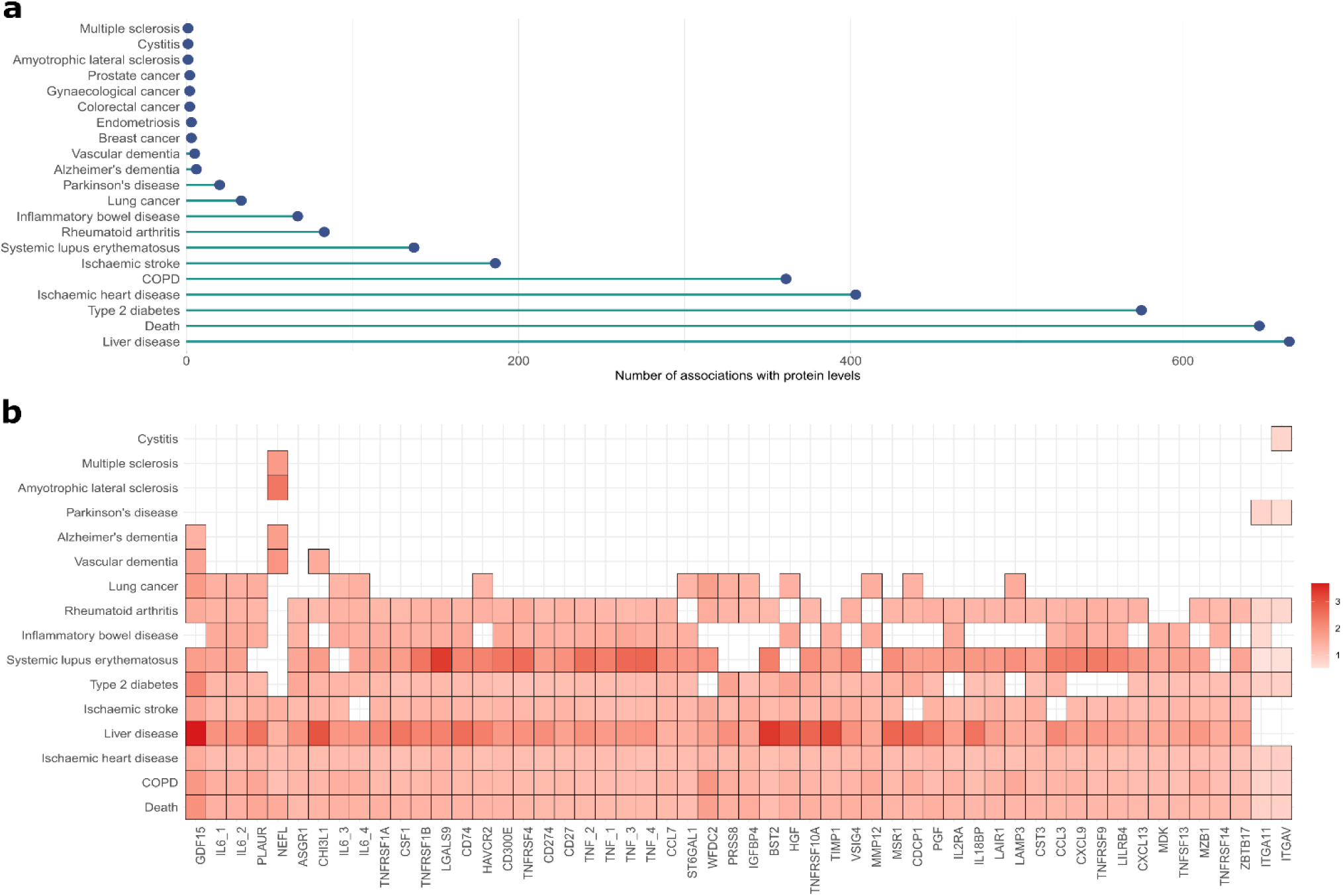
Individual protein associations with incident outcomes in the UK Biobank (N=47,600). **a,** Number of associations between protein analytes and time-to-onset for 20 outcomes that had P < 3.1×10^−6^ (Bonferroni-adjusted threshold) in both basic and fully-adjusted Cox PH models. There were 3,201 associations in total involving 961 protein analytes. **b,** Hazard ratios (HR) per a one SD increase in levels of the transformed protein analytes are plotted for the 54 protein analytes that were associated with eight or more outcomes in the individual Cox PH models. Each association is represented by a rectangle. Cox PH models were adjusted for age, sex and six lifestyle factors (BMI, alcohol consumption, social deprivation, educational attainment, smoking status and physical activity). Every association identified for these proteins had HR > 1 (red) and associations are shaded based on HR effect size (darkest colouration indicating larger magnitude of effect). The largest HR shown is for the association between GDF15 levels and liver disease (HR =3.67). COPD: chronic obstructive pulmonary disease.

### Proteomic signatures of multimorbidity

Fifty-four proteins had associations with eight or more incident morbidities **(Fig. 2b);** in all instances, elevated levels of the proteins were associated with the increased incidence of disease or death (i.e. HR > 1). Of the 54 proteins, GDF15 had the largest number of associations (11 incident outcomes), followed by IL6 and PLAUR (10 incident outcomes). In logistic regression models run between the 1,468 protein analytes and multimorbidity status (a binary trait defined as individuals that had three or more diagnoses of the 23 diseases over the 16-year follow-up period), 720 associations had P < 3.1×10^−6^ (**Supplementary Table 7)**. All 54 proteins that were associated with eight or more morbidities in the Cox PH associations were present in the multimorbidity status associations. GDF15, TNFRSF10B, WFDC2 and PLAUR had both the largest absolute effect sizes and smallest p-values, which was consistent with their position as top markers of multimorbidity in the individual Cox PH associations presented in **Fig. 2b**.

### Cox PH sensitivity analyses

Understanding whether protein-disease associations are stronger in the near-term of case follow-up is of interest when considering the clinical use-case for biomarkers. Modelling near-term versus long-term case follow-up is also important to understand the confidence that can be ascribed to associations failing the Cox PH assumption (Schoenfeld residual test P < 0.05). Therefore, a sensitivity analyses that modelled each of the 35,232 Cox PH associations over increasing yearly case follow-up intervals was performed **(Supplementary Table 8)**. Of the 3,201 protein-disease associations identified over the maximum 16-year follow-up, 2,915 and 1,957 of these associations remained (P < 3.1×10^−6^, the Bonferroni-adjusted threshold) when restricting cases up to 10-year and 5-year onset, respectively **(Supplementary Table 9)**. Of the 684 failures in the local (protein) Cox PH assumption observed in the 16-year follow-up analyses, 665 and 410 were observed in the 10-year and 5-year onset analyses. Relatively minor deviations in magnitude of effect size were observed for these associations by year of follow-up. These results can be examined visually for each of the 35,232 protein-disease associations tested in a Shiny app available at: https://protein-disease-ukb.optima-health.technology [Username: ukb_diseases, Password: UKBshinyapp]. The app also includes an interactive network for the 3,201 associations that can be manipulated to view multiple proteins and examine their associations with multiple incident morbidities.

A sensitivity analysis was performed to explore the potential impact of medication use on individual Cox PH associations. A subset of the population with proteomics measures had medication information available (35,073 of 47,600 individuals). Ischaemic heart disease was chosen, as a range of blood-pressure lowering medications are used to delay or prevent this disease and these medications were amongst the most commonly-reported in the population (14,074 of 35,073 individuals reported use of either statins, antihyperintensives, diuretics, beta blockers, calcium channel blockers or renin-angiotensin system actors at baseline **(Supplementary Table 10)**. In the subset of 35,073 individuals, 370 of the original 403 associations (adjusting for age, sex and six lifestyle factors) for ischaemic heart disease had P < 3.1×10^−6^ **(Supplementary Table 11).** With further adjustment for blood-pressure lowering medication use, 36 of these associations were attenuated, while 344 had P < 3.1×10^−6^. None of the attenuated associations were present in the top 100 marker associations (ranked by P-value, or effect size). Adjustment for blood-pressure lowering medication tended to reduce the magnitude of the effect estimate generally across the 370 associations **(Supplementary Fig.2)**, but hazard ratios were nonetheless highly correlated (Pearson correlation *r* = 0.99).

### ProteinScore development

ProteinScores for 19 diseases that had a minimum of 150 incident cases available were trained using Cox PH elastic net regression with cross-validation in a training subset. Cumulative time-to-onset distributions for cases **(Supplementary Figs. 3-4)** indicated that amyotrophic lateral sclerosis, endometriosis and cystitis were better-suited to 5-year onset assessments (80% of cases for these traits were diagnosed by year 8 of follow-up). All remaining ProteinScores were tested in the context of 10-year onset. Performance was quantified via incremental Cox PH models in the test subset, to obtain onset probabilities for calculation of AUC and Precision Recall AUC (PRAUC) estimates **(see Methods)**. This approach was repeated with fifty randomly sampled train and test subset combinations for each outcome to assess stability of ProteinScore performance given varied combinations of individuals in train and test sets. ProteinScores with the median difference in AUC beyond a minimally-adjusted model were selected for each outcome **(Supplementary Table 12)**. Summaries of protein features selected for the 19 ProteinScores are available in **Supplementary Tables 13-14**, ranging from five features selected for endometriosis to 143 features selected for type 2 diabetes **(Fig.3a)**.

**Figure 3.**
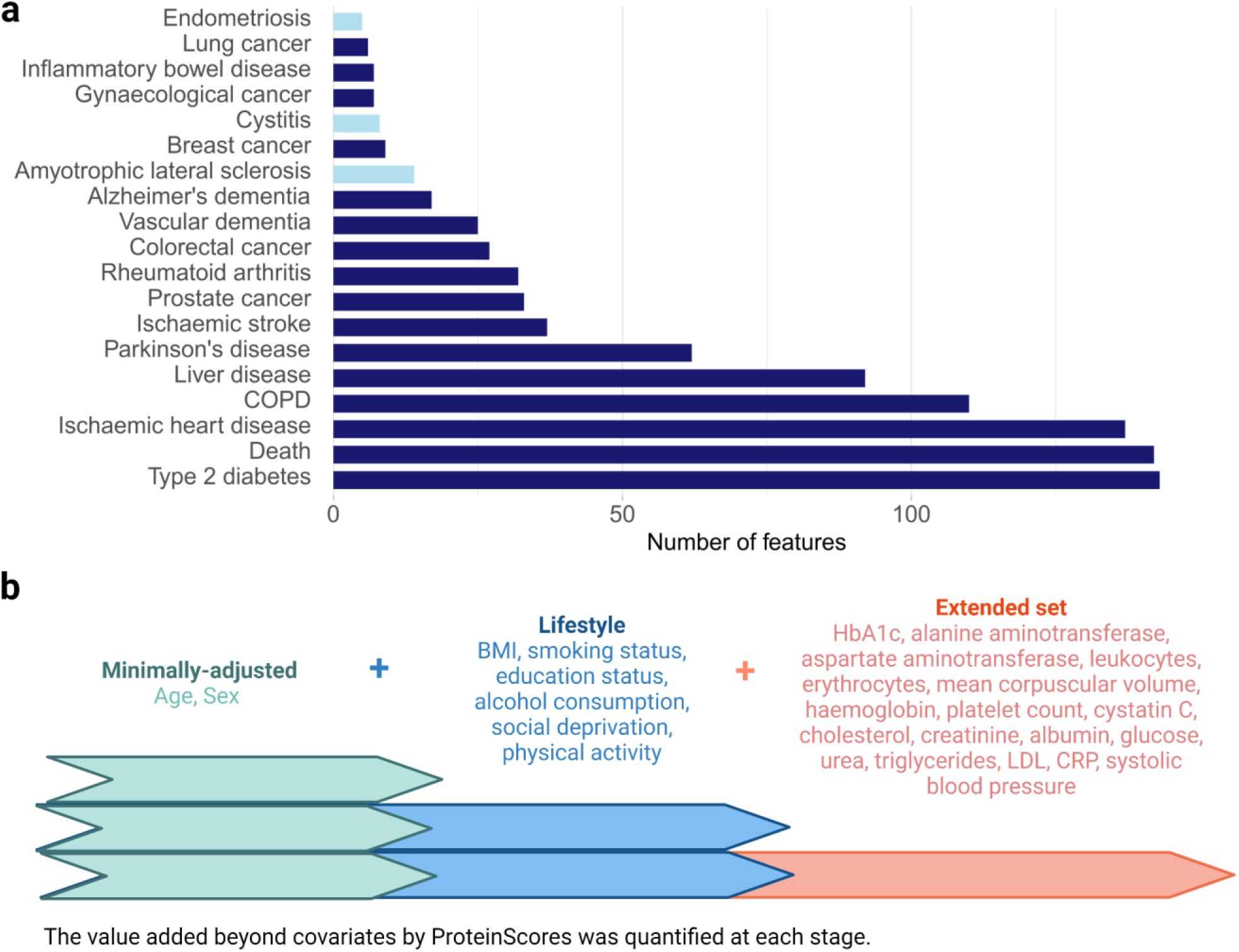
Feature selection and covariate structure for ProteinScore assessment. **a,** The total number of contributing protein analyte features selected for each ProteinScore. Incident outcomes that were assessed for 5-year onset (light blue) and 10-year onset (dark blue) are delineated. **b,** The increasingly comprehensive sets of covariates that were modelled in incremental Cox PH models to evaluate the value added by the ProteinScores beyond these covariates. These included an additional six lifestyle factors and 18 clinically-relevant biochemistry and physical measures. When modelled alongside age and sex, 26 possible covariates were therefore used in maximally-adjusted models.

### ProteinScore evaluation

Selected ProteinScores were evaluated alongside various combinations of covariates to quantify the additional improvements in AUC and PRAUC achieved by each score beyond these factors **(Fig.3b)**. Three increasingly complex sets of covariates were considered: 1) age and sex (where traits had not been sex-stratified), 2) further adjustment for a core set of six lifestyle and health covariates (BMI, alcohol consumption, social deprivation, educational attainment, smoking status and physical activity) and 3) further adjustment for an extended set of 18 biochemistry and physical attributes that are measurable in clinical settings. Performance when using only the ProteinScores was also considered, to ascertain whether protein information can streamline the signal offered by the set of 26 possible covariates. As these covariates are sourced from a range of physical measures, clinically-used biomarker assays and self-reporting, they represent a labour and time intensive resource that is rarely collated for every individual in clinical practice. A tabular summary of both the AUC and PRAUC statistics for all covariate combinations tested, with ROC P value comparisons comparing models with/out the addition of the ProteinScore are available in **Supplementary Table 15**. Strikingly, the singular inclusion of the ProteinScores had either equal or higher performance (as measured by AUC and PRAUC) than the maximal set of 26 covariates in eight instances (type 2 diabetes, liver disease, COPD, amyotrophic lateral sclerosis, death, Alzheimer’s dementia, ischaemic heart disease and Parkinson’s disease). The difference in AUC resulting from the addition of the ProteinScores into the three models with increasingly complex sets of covariates are summarised in **Fig.4a**.

**Figure 4.**
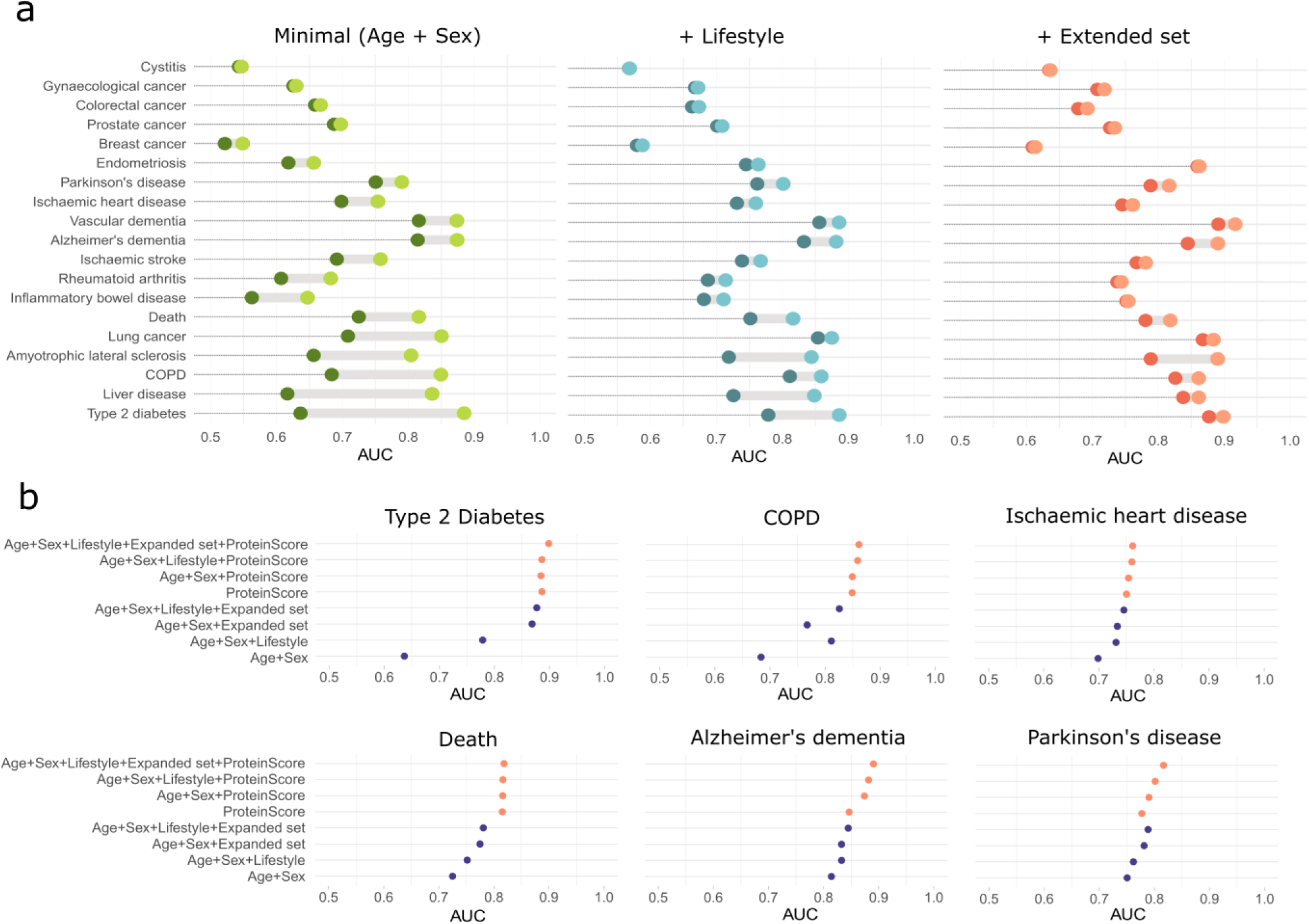
Predictive value offered by ProteinScores for incident outcomes in the UK Biobank. **a,** Differences in AUC resulting from the addition of the 19 ProteinScores to models with increasingly extensive sets of covariates: 1) minimally-adjusted (age and sex where traits were not sex-stratified), 2) minimally-adjusted with the addition of a core set of six lifestyle covariates and 3) further adjustment for an extended set of 18 covariates that are measured in clinical settings (physical and biochemical measures). AUC plots are ordered by increasing AUC differences in the minimally-adjusted models. All ProteinScore performance statistics shown correspond to 10-year onset, except those for ALS, endometriosis and cystitis that were assessed for 5-year onset. **b,** A breakdown of the AUC values achieved by different combinations of risk factors with/out the ProteinScores are shown for the six incident outcomes whereby the ProteinScore contributed statistically significant beyond a model including all 24 minimal, lifestyle and extended set variables (ROC P < 0.0026, the Bonferroni-adjusted threshold). All six of the best-performing ProteinScores shown were assessed for 10-year onset of disease. ALS: amyotrophic lateral sclerosis. COPD: chronic obstructive pulmonary disease.

In tests for significant differences between receiver operating characteristic (ROC) curves for the three models with increasingly complex sets of covariates with/out the ProteinScores, 10 of the ProteinScores (type 2 diabetes, liver disease, COPD, lung cancer, death, ischaemic stroke, Alzheimer’s and vascular dementias, ischaemic heart disease and Parkinson’s disease) had ROC P < 0.0026 (the Bonferroni-adjusted P-value threshold) beyond minimally-adjusted covariates. When adding ProteinScores to models that included both minimally-adjusted and lifestyle covariates, performance of lung cancer and vascular dementia ProteinScores was attenuated, leaving eight ProteinScores that had P < 0.0026 in ROC model comparison tests (type 2 diabetes, liver disease, COPD, death, ischaemic stroke, Alzheimer’s dementia, ischaemic heart disease and Parkinson’s disease). When assessing models that further adjusted for an additional 18 clinically-measurable covariates, six of the eight ProteinScores had P < 0.0026 in model comparisons with/out the ProteinScore, whereas liver disease and ischaemic stroke were attenuated by the extended covariate set. **Fig.4b** shows the breakdown of incremental model performance (by AUC) for each of these six ProteinScores: type 2 diabetes, COPD, death, Alzheimer’s dementia, ischaemic heart disease and Parkinson’s disease. Models that included only the ProteinScore are also presented with corresponding AUC performance.

### Exploration of the type 2 diabetes ProteinScore

To highlight the value that the best-performing ProteinScores may offer, type 2 diabetes was chosen as a case study for further exploration. Performance of the ProteinScore was assessed in the context of the current clinically-used biomarker for type 2 diabetes – glycated haemoglobin (HbA1c), in addition the predictive signals offered by other omics sources (genetic and metabolomic).

Given that the ProteinScore for type 2 diabetes added value beyond the extended set of covariates that included the well-validated biomarker HbA1c, the performance of HbA1c and the ProteinScore was directly compared in the test sample. As polygenic risk scores are also widely used to quantify the genetic risk contribution to disease, a polygenic risk score (PRS) for type 2 diabetes was also evaluated. In the type 2 diabetes test set, 1,105 cases (with mean time to onset 5.4 years [SD 3.0]) and 3,264 controls had all three measures available. HbA1c averages long-term glucose over two to three months and is widely employed clinically to monitor pre-clinical diabetes risk (42-47mmol/mol) and diagnose the disease (with two repeated measurements >48mmol/mol) ^29,30^. The rank-base inverse normal transformed levels of the ProteinScore and HbA1c had Pearson *r*=0.50 and discriminated incident case and control distributions similarly (**Fig. 5a**). HbA1c levels increased across ProteinScore risk deciles, with individuals in the upper deciles of the ProteinScore falling within the clinical HbA1c screening threshold (42-47mmol/mol) for diabetes **(Fig. 5b)**. In incremental Cox PH models for the 10-year onset of type 2 diabetes **(Fig. 5c)** the singular use of the ProteinScore (AUC = 0.89) outperformed both HbA1c (AUC = 0.85) and the PRS (AUC = 0.68). In ROC model comparisons between HbA1c alone and HbA1c with the ProteinScore added, a statistically significant improvement due to the ProteinScore was identified (ROC P < 0.0026). When the PRS was added to this model (including HbA1c and the ProteinScore), AUC remained unchanged (0.91), whereas PRAUC improved (from 0.79 to 0.80) and a significant difference due to the addition of the PRS was identified (ROC P < 0.0026). **Supplementary Table 16** summarises the results from these analyses, which are also presented in **Fig.5c**.

**Figure 5.**
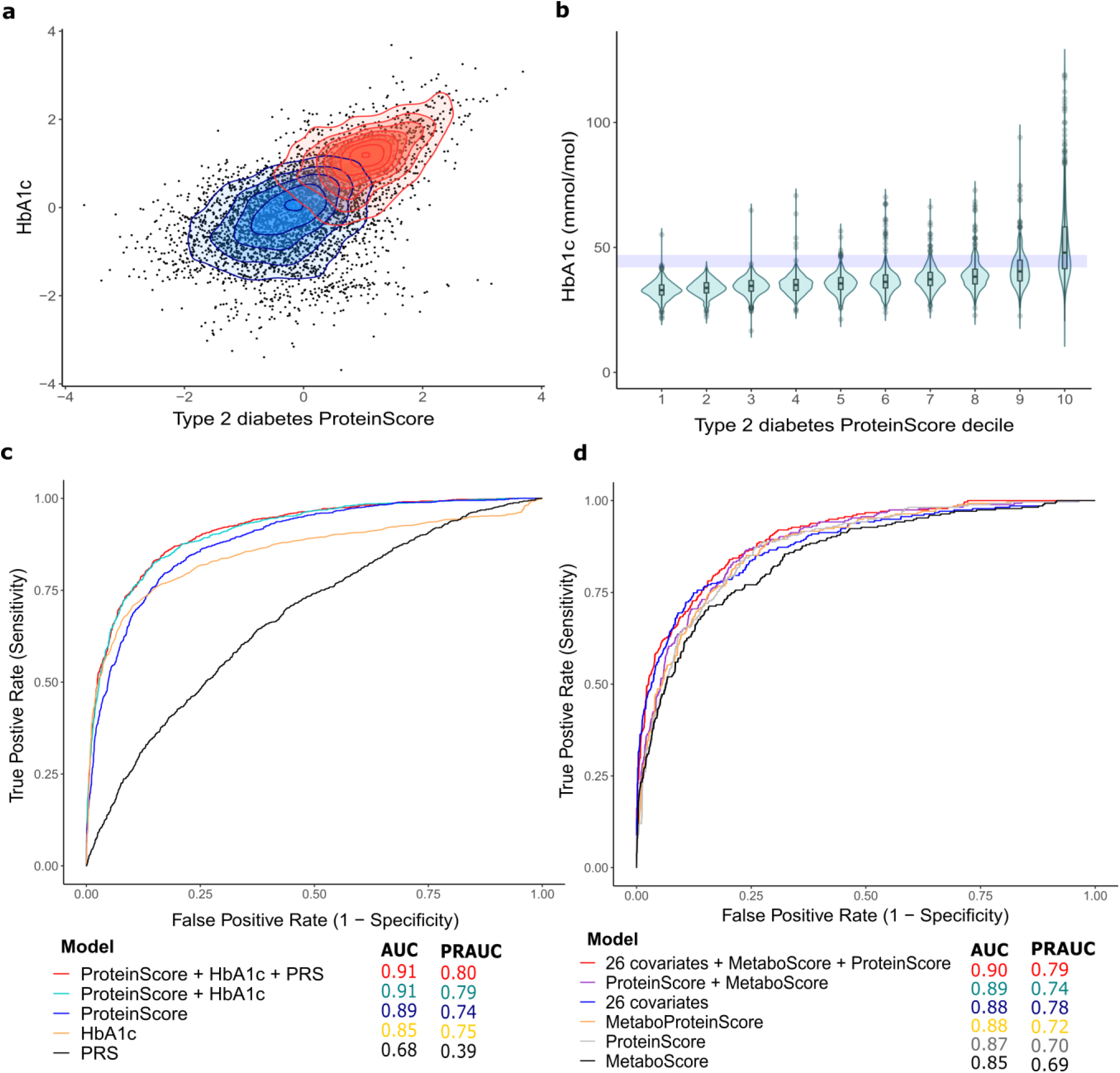
Exploration of the type 2 diabetes ProteinScore. **a,** Case (red) and control (blue) discrimination for HbA1c and the type 2 diabetes ProteinScore in the test set (1,105 cases, 3,264 controls, mean time to case onset 5.4 years [SD 3.0]). Both markers were rank-based inverse normalised and scaled to have a mean of 0 and standard deviation of 1. **b,** HbA1c (mmol/mol) per decile of the type 2 diabetes ProteinScore in the test set. The shaded rectangle indicates the type 2 diabetes HbA1c screening threshold (42-47 mmol/mol). **c,** ROC curves for incremental 10-year onset models incorporating HbA1c, the type 2 diabetes ProteinScore and a polygenic risk score (PRS) for type 2 diabetes individually and concurrently. **d,** ROC curves for 10-year onset scores developed in the subsets of the type 2 diabetes train and test populations that had metabolomics and proteomics available (N cases_train_ = 377, N controls_train_ = 1,002, N cases_test_ = 309, N controls_test_ = 898). A Metabolomic score (MetaboScore), ProteinScore, and a joint omics score (MetaboProteinScore) are modelled individually and concurrently and benchmarked against 26 covariates (age, sex, six lifestyle factors and the extended set of 18 clinically-relevant covariates.

In a preliminary assessment, we aimed to 1) directly compare scores generated with either metabolomics-only or protein-only features and 2) assess the value added through the consideration of proteomic and metabolomic features simultaneously. The original type 2 diabetes ProteinScore populations were subset to training and testing sets that had proteomic and metabolomic measures available (N cases_train_ = 377, N controls_train_ = 1,002, N cases_test_ = 309, N controls_test_ = 898). Performance of a MetaboScore (considering metabolite features), a ProteinScore (considering protein features) and a MetaboProteinScore (combining metabolomic proteomic features) is summarised for the restricted population in **Fig. 5d**. The ProteinScore (AUC = 0.87) outperformed the MetaboScore (AUC = 0.85) The MetaboProteinScore (that considered both omics measures as potential features) had an AUC of 0.88, whereas modelling the independently-trained MetaboScore and ProteinScore together resulted in an AUC of 0.89. The maximal set of 26 possible covariates **(see Fig.3b)** had an AUC of 0.88, which rose to a maximal AUC of 0.90 upon the inclusion of the MetaboScore and ProteinScore. The selected features and weights for each score (MetaboScore = 19 features, ProteinScore = 52 features and MetaboProteinScore = 37 features) are available in **Supplementary Table 17**, with full AUC and PRAUC statistics available in **Supplementary Table 18**.

## Discussion

Identifying individuals at risk of a future disease event or death is a priority for prevention-based medicine during ageing ^31^. We report 3,201 associations between 961 circulating proteins and 21 incident outcomes, identifying proteins indicative of multimorbidity. ProteinScores for incident type 2 diabetes, COPD, ischaemic heart disease, Alzheimer’s dementia, Parkinson’s disease and death demonstrated value beyond a comprehensive set of 26 covariates, offering comparable performance and minimising the need for extensive recording of lifestyle factors, physical measures and biomarker assays. Exploration of the type 2 diabetes ProteinScore suggested that while protein information captures much of the predictive signal, augmenting traditional risk factors with proteomic, metabolomic and genetic data types may further hone risk classification.

The breadth of electronic health data linkage and protein data available in UK Biobank provides a unique resource for profiling early molecular signatures of age-related disease. This study demonstrates that for certain diseases, subsets of relatively few circulating proteins can add predictive value, up to a decade prior to formal diagnoses. As available cases increase, it is likely that the performance of ProteinScores will be enhanced. Nonetheless, for the best-performing six ProteinScores, modelling the ProteinScore in isolation resulted in equal or higher AUCs than models with extensive covariate adjustments. This suggests that ProteinScores for such traits absorb a large proportion – if not all – of the signal and may offer a streamlined set of metrics to proxy for an individual’s health status. This often-enhanced predictive quality of the scores presents an exciting opportunity to reconsider how best to formulate (and maintain) modern clinical prediction models. This is an important consideration given that self-reported measures are known to be variable in accuracy and are often misreported ^32^. Additionally, while much interest is currently devoted to employing PRS for disease prediction, they neglect environmental components of disease risk and may therefore be limited in the context of complex age-related disease ^33,34^. Our ProteinScore for type 2 diabetes outperformed the PRS, which is likely due to proteins representing an interface that captures genetic, environmental and lifestyle contributions to disease risk. The improvement in AUC resulting from concurrent modelling of HbA1c and the type 2 diabetes ProteinScore suggests that the latter may provide additional predictive value. Similarly, in a subset of the population with metabolomics measures, the type 2 diabetes ProteinScore outperformed the MetaboScore, with an additive signal achieved by modelling both scores. While diabetes is typically considered to be a metabolic disease, the breadth of coverage (249 metabolites, versus 1,468 protein measures) may limit the metabolic score performance. ProteinScores for multiple diseases within the same individuals may facilitate an improved understanding of multimorbidity. For example, if an individual falls within the top 5% of the ProteinScore distributions for type 2 diabetes and Alzheimer’s dementia, this information may enhance personalised intervention plans. The ProteinScore performance for Alzheimer’s dementia was also largely unchanged upon addition of additional covariates. As therapeutic interventions for neurodegenerative diseases have greater efficacy when implemented earlier in the disease pathogenesis ^35–37^, the ProteinScore for Alzheimer’s dementia may hone trial recruitment.

The method for ProteinScore generation selects proteins that, in combination, are predictive of outcomes, but these do not necessarily represent the most probable drivers of disease. It is likely that a subset of the 3,201 individual protein-disease associations we report represent direct mediators of disease. The goal of this work was to identify early markers that associate with incident disease and the markers we identify are therefore useful for risk stratification purposes (even if they are indicative of underlying morbidities at baseline, or do not represent causal mediators). To delineate the markers that may be direct mediators of disease, we encourage further exploration of through techniques such as Mendelian randomisation and colocalisation. Similarly, further modelling that takes into account multimorbidity trajectories over the lifecourse would also aid in understanding the role of prevalent diseases and medication use on future disease risk. The largest number of associations and strongest effect sizes (by magnitude of the absolute log of the hazard ratio) were observed for liver disease in individual Cox PH analyses. For neurological diseases and cancers, where fewer associations were identified, it is possible that the blood is less able to capture the full spectrum of disease pathogenesis, which may be localised to distal tissues. Similarly, the panel of proteins available may reflect certain diseases better than others.

All 54 proteins that were associated with eight or more morbidities had associations with hazard ratios greater than 1, indicating that elevated levels of these proteins may serve as early warning signatures of disease onset. Elevated growth differentiation factor 15 (GDF15), Interleukin-6 (IL6) and plasminogen activator urokinase receptor (PLAUR) had the largest number of associations with incident diseases. This result is in concordance with previous screening of the circulating proteome against multimorbidity and mortality, which identified GDF15 as the top marker of future multimorbidity from 1,301 plasma proteins tested ^38,39^. Further evidence supporting GDF15 as a marker of multiple outcomes including heart disease, type 2 diabetes, stroke, dementia and death has been reported ^39–45^. IL6 mediates chronic, low-grade inflammation, is a key biomarker of ageing ^46^ and anti-IL6 therapeutics have been developed for a range of inflammation-associated diseases ^47,48^. While less-extensive evidence exists supporting PLAUR as a biomarker of multiple morbidities, it was associated with incident cancer, cardiovascular disease, diabetes and mortality in previous Cox PH analyses ^49^. Similarly, increased levels of neurofilament light (NEFL) were associated with higher incidence of multiple neurological traits (Parkinson’s disease, Alzheimer’s dementia, multiple sclerosis, amyotrophic lateral sclerosis and ischaemic stroke). These diseases are hallmarked by neuron degradation and NEFL may therefore be a consequential marker that is released into the blood upon breakdown of synapses ^50,51^. NEFL was also associated with liver disease, COPD and ischaemic heart disease, which may reflect the presence of underlying synaptic and neuronal dysfunction, or the presence of comorbidities in individuals with these diagnoses.

Across the 16-year window of follow-up in individual Cox PH models, a subset of associations violated the Cox PH assumption at the local (protein) level. Our Shiny app https://protein-disease-ukb.optima-health.technology [Username: ukb_diseases, Password: UKBshinyapp] provides visualisations for sensitivity analyses run across cases over successive years of case follow up, allowing for interrogation of the stability of individual protein-disease relationships. This information on near-term versus long-term case follow-up is often of importance to clinicians and patients for behaviour change and intervention strategies. The Shiny app also visualises the 3,201 fully-adjusted associations in a network view, allowing users to view overlapping signatures between multiple proteins and the onset of multiple diseases.

This study has several limitations. First, a subset of 6,385 individuals in the UKB-PPP sample were selected by consortium members for enrichment of certain diagnoses and this non-random selection can introduce biases. Second, as UK Biobank currently represents the largest population with comprehensive Olink proteomics and electronic health data linkage, it was not possible to source an external test set for the ProteinScores. Third, variation in protein analyte levels across measurement technologies has been reported ^52^. Results should therefore be corroborated across panels in future. Fourth, the protein measured were recorded in relative scale, rather than absolute quantification. This limits direct translation of the ProteinScores for direct prediction in new populations. However, the early markers of incident disease that are identified may still replicate when datasets become available to facilitate replication analyses. Fifth, the UK Biobank population is largely comprised of individuals with European ancestry and a restricted age range (40-71 years, with a mean of 57 years); future studies in equally well-characterized cohorts will be needed to assess how well ProteinScores translate to other populations and ethnicities. Sixth, non-linear trajectories of blood-based protein signatures are known to exist across the life course in the context of ageing ^53^. These factors should be considered in disease-specific analyses in future. Seventh, death was treated as a censoring event; competing risks and multi-state modelling approaches may be used for disease-protein associations in future to resolve the impact of death as a competing risk for disease onset. Finally, although a comprehensive set of major age-related morbidities were studied, many diseases were not included in this work. Continued linkage and proteomic sampling will expand the applications of ProteinScores to further diseases.

In conclusion, this study quantified circulating proteome signatures that are reflective of multiple individual disease states across mid-to-later life. ProteinScores for the incidence of six incident outcomes significantly improved AUCs for 10-year onset beyond 26 demographic, lifestyle and clinically-relevant covariates. The type 2 diabetes ProteinScore offered additional value beyond HbA1c, a PRS and a metabolomic score. A total of 3,123 individual protein-disease associations were also profiled across the 16-year follow-up period, identifying candidate targets for multimorbidity prevention. These data suggest that proteomic features are powerful tools for honing risk stratification.

## Methods

### The UK Biobank sample population

UK Biobank (UKB) is a population-based cohort of around 500,000 individuals aged between 40-69 years that were recruited between 2006 and 2010. Genome-wide genotyping, exome sequencing, electronic health record linkage, whole-body magnetic resonance imaging, blood and urine biomarkers and physical and anthropometric measurements are available. More information regarding the full measurements can be found at: https://biobank.ndph.ox.ac.uk/showcase/. The UK Biobank Pharma Proteomics Project (UKB-PPP) is a precompetitive consortium of 13 biopharmaceutical companies funding the generation of blood-based proteomic data from UKB volunteer samples.

### Proteomics in the UK Biobank

The UKB-PPP sample includes 54,306 UKB participants and 1,474 protein analytes measured across four Olink panels (Cardiometabolic, Inflammation, Neurology and Oncology: annotation information provided in **Supplementary Table 1**) ^25^. A randomised subset of 46,673 individuals were selected from baseline UKB, with 6,385 individuals selected by the UKB-PPP consortium members and 1,268 individuals included that participated in a COVID-19 study. The randomised samples have been shown to be highly representative of the wider UKB population, whereas the consortium-selected individuals were enriched for 122 diseases ^25^. Details on sample selection for UKB-PPP, in addition to processing and quality control information for the Olink assay are provided in **Supplementary Information**. Of 54,309 individuals that had protein data measured, there were 52,744 that were available after quality control exclusions with 1,474 Olink protein analytes measured (annotations in **Supplementary Table 1**) ^25^. The sample is predominantly white/European (93%), but also has individuals with black/black British, Asian/Asian British, Chinese, mixed, other and missing ethnic backgrounds (7%).

**Supplementary Fig. 1** summarises the processing steps applied to this dataset to derive a complete set of measurements for use. Briefly, of 107,161 related pairs of individuals (calculated through kinship coefficients > 0 across the full UKB cohort), 1,276 pairs were present in the 52,744 individuals. After exclusion of 104 individuals in multiple related pairs, in addition to one individual randomly selected from each of the remaining pairs, there were 51,562 individuals. A further 3,962 individuals were excluded due to having >10% missing protein measurements. Four proteins that had >10% missing measurements (CTSS.P25774.OID21056.v1 and NPM1.P06748.OID20961.v1 from the neurology panel, PCOLCE.Q15113.OID20384.v1 from the cardiometabolic panel and TACSTD2.P09758.OID21447.v1 from the oncology panel) were then excluded. The remaining 1% of missing protein measurements were imputed by K-nearest neighbour (k=10) imputation using the impute R package (Version 1.60.0) ^54^. The final dataset consisted of 47,600 individuals and 1,468 protein analytes. Assessments of protein batch, study centre and genetic principal components suggested that these factors had minimal effects on protein levels (lowest correlation between protein levels and residuals of 0.94) **(Supplementary Information)**. Therefore, protein levels were not adjusted for these factors.

### Phenotypes in the UK Biobank

Demographic and phenotypic information for the 47,600 individuals with complete protein data for 1,468 analytes are available in **Supplementary Table 2**. Lifestyle covariates included: BMI (weight in kilograms divided by height in metres squared), alcohol intake frequency (1 = Daily or almost daily, 2 = Three-Four times a week, 3 = Once or twice a week, 4 = One-Three times a month, 5 = Special occasions only, 6 = Never), the Townsend index of deprivation (higher score representing greater levels of deprivation) and smoking status (0 = Never, 1 = Previous, 2 = Current), physical activity (0 = between 0-2 days/week of moderate physical activity, 1 = between 3-4 days/week of moderate physical activity, 2 = between 5-7 days/week of moderate physical activity) and education status (1 = college/university educated, 0 = all other education). Of the 47,600 individuals with complete protein data, there were 52, 52, 236, 56 and 59 missing entries for alcohol, smoking, BMI, physical activity and deprivation, respectively. No imputation of missing data was performed for the inclusion of these variables in individual Cox PH analyses. There were an additional 2,556, 188 and 59 individuals that answered ‘prefer not to answer’ and were excluded from physical activity, smoking and alcohol variables, respectively.

### Electronic health data linkage in the UK Biobank

Electronic health linkage to NHS records was used to collate incident diagnoses. Death information was sourced from the death registry data available through the UK Biobank. Cancer outcomes were sourced from the cancer registry (ICD codes), whereas non-cancer diseases were sourced from first occurrence traits available in the UK Biobank. The first occurrence traits integrate GP (read2/3), ICD (9/10) with self-report and ICD codes present on the death registry to identify the earliest date of diagnoses. These data sources are linked to 3-digit ICD trait codes. A summary of codes used to extract each of the outcomes included in the present study are detailed in **Supplementary Information**. The following 23 diseases were included: liver disease, systemic lupus erythematosus, type 2 diabetes, amyotrophic lateral sclerosis, Alzheimer’s dementia, endometriosis, chronic obstructive pulmonary disease (COPD), inflammatory bowel disease, rheumatoid arthritis, ischaemic stroke, Parkinson’s disease, vascular dementia, ischaemic heart disease, major depressive disorder, schizophrenia, multiple sclerosis, cystitis and lung, prostate, breast, gynaecological, brain/CNS and colorectal cancers. These represent a selection of leading age-related causes of morbidity, mortality and disability. In all analyses involving sex-specific diseases, the population was stratified to males or females and sex was not included as a covariate in incremental Cox PH assessments. Traits that were stratified included gynaecological cancer, breast cancer, endometriosis and cystitis (all female-stratified) and prostate cancer (male-stratified).

### Incident disease calculation in the UK Biobank

Dates of diagnoses for each disease were ascertained through electronic health linkage. Using the date of baseline appointment, time-to-first-onset for each diagnoses in years was calculated. Time-to-onset for controls was defined as the time from baseline to censoring date (**Supplementary Information**). Death was treated as a censoring event. Time-to-censor date was calculated for the controls that remained alive, whereas if a control individual had died during follow-up time-to-death was taken forward for Cox PH models. Any cases that were prevalent at baseline were excluded. Alzheimer’s and vascular dementias were restricted to age at onset (or censoring) of 65 years or older in all analyses. Sex-specific traits were stratified across all analyses.

### Individual Cox proportional hazards analyses

Cox proportional hazards models were run between each protein and each incident disease using the ‘survival’ package (Version 3.4-0) ^55^ in R (Version 4.2.0) ^56^. Protein levels were rank-based inverse normalised and scaled to have a mean of 0 and standard deviation of 1 prior to analyses. Minimally-adjusted Cox PH models for sex-stratified traits included age at baseline as a covariate, whereas the remaining models adjusted for age and sex. Lifestyle-adjusted models further controlled for education status, BMI, smoking status, social deprivation rank, physical activity and alcohol intake frequency. A Bonferroni-adjusted P-value threshold for multiple testing based on the 678 components that explained 90% of the cumulative variance in the 1,468 protein analyte levels **(Supplementary Table 3)** and 24 outcomes tested was applied across all Cox PH models (P < 0.05/(678 × 24) = 3.1×10^−6^ used as the Bonferroni-adjusted P-value threshold). Proportional hazards assumptions were checked through examination of protein-level Schoenfeld residuals.

A sensitivity analysis was performed for each of the 35,232 fully-adjusted associations tested, restricting cases to successive years of follow-up. These sensitivity analyses were visualised using the Shiny package (Version 1.7.3) ^57^ in R. The magnitude of change in hazard ratios for individual associations can be examined by year of case follow-up to assess consistency of effect sizes. Whether marker associations are stronger or weaker when restricting to cases occurring in the near-term (1-5 years of follow-up) can also be examined. A network visualisation was also created within the Shiny interface to highlight the fully-adjusted associations that had P < 3.1×10^−6^ using networkD3 (Version 3.0.4) ^58^ and igraph (Version 1.3.5) ^59^ R packages. To further verify the markers of multiple morbidities identified in individual Cox PH analyses, logistic regression models were also run between each of the 1,468 protein analyte levels and multimorbidity status (defined as 1,454 individuals that received 3 or more of the 23 disease diagnoses over the 16-year follow-up period). A sensitivity analyses was also run for ischaemic heart disease associations with/out adjustment for blood-pressure lowering medication reported at baseline in a subset of individuals (35,073 of 47,600) that had medication information available. **Supplementary Information** provides details on the classification of medications as per the anatomical therapeutic chemical (ATC) classification categories. A total of 14,074 individuals (of the 35,073) indicated they were taking one or more of the above blood-pressure lowering medications at baseline. This was treated as a binary variable and the comparison with/out adjustment for this variable was performed for ischaemic heart disease Cox PH associations in the subset of 35,073 individuals. Adjustments for age, sex and six lifestyle factors were included in both sets of analyses, with 2,456 cases, 27,468 controls.

### ProteinScore development

MethylPipeR ^60^ is an R package with accompanying user interface that we have previously developed for systematic and reproducible development of incident disease predictors. Using MethylPipeR, ProteinScores that considered 1,468 Olink protein levels were trained using Cox PH elastic net regression via the R package Glmnet (Version 4.1-4) ^61^. Penalised regression minimises overfitting by the use of a regularisation penalty and the best shrinkage parameter (λ) was chosen by cross-fold validation with alpha fixed to 0.5. Of the 24 outcomes featured in the individual Cox PH analyses, 19 that had a minimum case count of 150 were selected for ProteinScore development. The chosen strategy for ProteinScore development included training ProteinScores for each trait across fifty randomised iterations (with each iteration including a different combination of cases and controls in train and test sets). This strategy quantifies the stability of the ProteinScore performance, which is critical given that unobserved confounders that may be enriched during random selection of individuals from the wider population. The ProteinScore training strategy is summarised in **Supplementary Fig. 5**. Briefly, 50 iterations of each ProteinScore were performed that randomised sample selection by 50 randomly sampled seeds (values between 1 and 5000). For each iteration, cases and controls were randomly split into 50% groups for training and testing. From the 50% training control population, a subset of controls were then randomly sampled to give a case:control ratio of 1:3 in order to balance the datasets. For traits with over 1000 cases in training samples 10 folds were used. For traits with between 500 and 1000 cases in training, five folds were used. Three folds were used when there were fewer than 500 cases in the training sample. Protein levels were rank-based inverse normalised and scaled to have a mean of 0 and standard deviation of 1 in the training set. The linear combination of weighting coefficients for selected protein features from cross-validation within the folds of the training set were then used to generate a ProteinScore for each individual in the test samples. Of the 50 training iterations tested, models that had no features selected were documented **(Supplementary Table 12)**.

### Assessment of ProteinScore performance

Cumulative time-to-onset distributions for cases **(Supplementary Figs. 3-4)** indicated that amyotrophic lateral sclerosis, endometriosis and cystitis were better-suited to 5-year onset assessments in the test sample (80% of cases were diagnosed at 8-years post-baseline). All remaining ProteinScores were tested in the context of 10-year onset (80% of cases were not diagnosed 8-years post-baseline). Across the 50 ProteinScore iterations for each trait, 50% of cases and controls that were not randomly selected for training were reserved for testing. For a visualisation of the test set sampling and assessment strategy, see **Supplementary Fig. 5**. In the test set, cases that had time-to-event up to or including the 5-year or 10-year thresholds used for onset prediction were selected, while cases beyond the threshold were placed with the control population, which was then randomly sampled in a 1:3 ratio. Weighting coefficients for features selected during ProteinScore training were used to project scores into the test sample. Incremental Cox PH models were run in the test sample to obtain cumulative baseline hazard and onset probabilities, which were used to derive AUC and PRAUC estimates. The test set sampling strategy ensured that while the majority of cases occurred up to the onset threshold, there were a small proportion (∼3%) of cases included in Cox PH models with onset times after the 10- or 5-year threshold, to simulate a real-world scenario for risk stratification. If cases fell beyond the 5-year or 10-year threshold for onset, they were recoded as controls in the AUC calculation. Cumulative baseline hazard probabilities were calculated using the Breslow estimator available in the ‘gbm’ R package (Version 2.1.8.1) ^62^. Survival probabilities were then generated through taking the exponential of the negative cumulative baseline hazard at 5 or 10 years to the power of the Cox PH prediction probabilities. ProteinScore onset probabilities were calculated as one minus these survival probabilities. AUC, PRAUC and ROC statistics were extracted for the survival probabilities using the calibration function from the ‘caret’ R package (Version 6.0-94) ^63^ and the evalmod function from the ‘MLmetrics’ R package (Version 1.1.1) ^64^.

ProteinScores that yielded the median incremental difference to the AUC of a minimally-adjusted model (adjusting for age- or age- and sex) were selected from the fifty possible ProteinScores for each trait. If no features were selected during training, models were weighted as performance of 0 in the median model selection. In some instances, features were selected during training and incremental Cox PH models were run successfully, but the random sampling of the test set did not include a case with time-to-event at or after the 5-year or 10-year onset threshold. Therefore, these models were excluded as cumulative baseline hazard distributions did not reach the onset threshold and could not be extracted for AUC and PRAUC calculations. The number of models, with minimum and maximum performance was documented **(Supplementary Table 12)**. Taking this approach mitigated against the presence of extreme case:control profiles driving ProteinScore performance and minimised the possibility of bias being introduced by selecting train and test samples based on matching for specific population characteristics.

Selected ProteinScores for each trait were then evaluated to quantify the additional value (in terms of increases in AUC and PRAUC) that resulted from the addition of ProteinScores. Minimally-adjusted models included age and sex (if traits were not sex-stratified). Lifestyle-adjusted models then further accounted for common lifestyle covariates (education status, BMI, smoking status, social deprivation rank, physical activity and alcohol intake frequency). Finally, models included covariates from the minimally-adjusted, lifestyle-adjusted and an extended set of clinically-measured variables were then assessed **(see Fig.3b)**. In each case, the difference in AUC and PRAUC resulting from the addition of the ProteinScore was reported. ROC P-value tests were used to ascertain whether the improvements offered by selected ProteinScores for each outcome were statistically significant, beyond each set of increasingly saturated covariates. A Bonferroni-adjusted P-value threshold for ROC P tests was used based on the 19 ProteinScore traits (P < 0.05/19 = 0.0026). The ‘precrec’R package (Version 0.12.9) ^65^ was used to generate ROC and Precision-Recall curves for each ProteinScore. A series of models that included only the ProteinScore were also considered for each outcome, to quantify whether protein data alone could absorb much of the predictive performance achieved by the covariates.

A set of 26 possible covariates used across the minimally-adjusted, lifestyle-adjusted and extended set analyses were assessed for missingness, imputed (where missingness was < 10%) and utilised in ProteinScore evaluation as a maximal, extended set of covariates. Further details of variable selection and preparation are supplied in **Supplementary Information**. Additional covariates (considered in addition to age, sex and six lifestyle traits that were used in individual Cox PH analyses) included: leukocyte counts (10^9 cells/Litre), erythrocyte counts (10^12 cells/Litre), haemoglobin concentration (grams/decilitre), mean corpuscular volume (femtolitres), platelet count (10^9 cells/Litre), cystatin C (mg/L), cholesterol (mmol/L), alanine aminotransferase (U/L), creatinine (umol/L), urea (mmol/L), triglycerides (mmol/L), LDL (mmol/L), CRP (mg/L), aspartate aminotransferase (U/L), glycated haemoglobin – HbA1c – (mmol/mol), albumin (g/L), glucose (mmol/L) and systolic blood pressure (mmHg). After covariate processing steps were complete, a population of 43,437 individuals was available with complete information for ProteinScore testing. Phenotypic summaries of the additional covariates for this population are sumamrised in **Supplementary Table 2**.

### Further assessment of the type 2 diabetes ProteinScore

Glycated Haemoglobin (HbA1c) is a blood-based measure of chronic glycemia that is highly predictive of type 2 diabetes events and is recommended as a test of choice for the monitoring and diagnosis of type 2 diabetes ^29,30^. HbA1c (mmol/mol) measurements (fieldID 30750) and the type 2 diabetes polygenic risk score (PRS) available in UK Biobank (fieldID 26285) were extracted. A contour plot showing both variables grouped by those who went on to be diagnosed with type 2 diabetes over a 10-year period was created. HbA1c levels were also plotted against ProteinScore risk deciles. HbA1c and the ProteinScore levels were rank-based inverse normalised and assessed individually and concurrently in incremental models for 10-year onset of type 2 diabetes in the ProteinScore test set. A Pearson correlation coefficient (*r*) between the transformed HbA1c and ProteinScore levels was calculated. The 10-year incremental Cox PH models were used to derive onset probabilities for calculation of AUCs and PRAUCs after adding the ProteinScore to models adjusting for HbA1c and the type 2 diabetes PRS. Model comparisons were used (test of the difference in ROC curves) to quantify the value added by the ProteinScore beyond the PRS and HbA1c.

Metabolomics measures were available in 12,050 of the 47,600 individuals with proteomic data included in the study (see **Supplementary Information** for details on data preparation). Type 2 diabetes was chosen as a case study for further exploration, as it is typically well-reflected by circulating levels of both protein and metabolomic markers and is considered as a metabolic disease. The train and test sets used to develop the main type 2 diabetes ProteinScore were subset to those with metabolomics available (N cases_train_ = 377, N controls_train_ = 1,002, N cases_test_ = 309, N controls_test_ = 898). Scores that considered only metabolomic features (MetaboScore), only proteomic features (ProteinScore) and joint omics features (MetaboProteinScore) were trained and tested in these populations. There were 249 metabolite levels and 1,468 protein levels considered as potentially-informative features. Performance was evaluated for 10-year onset of type 2 diabetes in the test sample, modelling the scores individually, concurrently and benchmarking them against the maximal set of 26 possible covariates **(see Fig.3)**.

### Ethics declarations

All participants provided informed consent. This research has been conducted using the UK Biobank Resource under approved application numbers 65851, 20361, 26041, 44257, 53639, 69804.

## Data availability

Datasets generated in this study are made available in **Supplementary Tables**. Proteomics data is available in the UK Biobank under Category 1838 at: https://biobank.ndph.ox.ac.uk/ukb/label.cgi?id=1838.

## Code availability

Code is available with open access at the following Github repository: https://github.com/DanniGadd/Blood_protein_levels_and_incident_disease_UK_Biobank

## Acknowledgements

**This research was funded in whole, or in part, by the Wellcome Trust [108890/Z/15/Z]. For the purpose of open access, the author has applied a CC BY public copyright licence to any Author Accepted Manuscript version arising from this submission.**

We thank the participants, contributors, and researchers of UK Biobank for making data available for this study – with special thanks to Lauren Carson, John Busby, Naomi Allen and Rory Collins for making the study possible. We are grateful to the research & development leadership teams at the thirteen participating UKB-PPP member companies (Alnylam Pharmaceuticals, Amgen, AstraZeneca, Biogen, Bristol-Myers Squibb, Calico, Genentech, Glaxo Smith Klein, Janssen Pharmaceuticals, Novo Nordisk, Pfizer, Regeneron, and Takeda) for funding the study. We thank the Legal and Business Development teams at each company for overseeing the contracting of this complex, precompetitive collaboration – with particular thanks to Erica Olson of Amgen, Andrew Walsh of GSK, and Fiona Middleton of AstraZeneca. The Biogen team is especially thankful to Helen McLaughlin for her project management support. Finally, we thank the team at Olink Proteomics (Philippa Pettingell, Klev Diamanti, Cindy Lawley, Linda Jung, Sara Ghalib, Ida Grundberg and Jon Heimer) for their consistent logistic support throughout the project – with special thanks to Evan Mills for co-championing the project and leading internal activities at Olink.

R.E.M. is supported by Alzheimer’s Society major project grant AS-PG-19b-010. R.F.H is supported by a MRC IEU Fellowship. D.A.G. is supported by the Wellcome Trust Translational Neuroscience programme [**108890/Z/15/Z]**.

## Author contributions

D.A.G., R.F.H., R.E.M., B.S., C.F., and Z.K., conceptualised the study design and consulted on methods and results. D.A.G., carried out all analyses. D.A.G., R.F.H., B.B.S., and R.E.M., drafted the article. R.A., and J.G., conducted preliminary analyses. T.L., and K.F., performed quality control on the proteomics dataset. Y.C., was consulted on methodology. M.D. contributed to Shiny app integration of results. All authors reviewed and approved of the manuscript.

## Materials and correspondence

Correspondence and material requests should be sent to Dr Benjamin Sun at or Prof Riccardo Marioni at.

