## Supplementary Figures for "Blood protein levels predict leading incident diseases and mortality in UK Biobank"

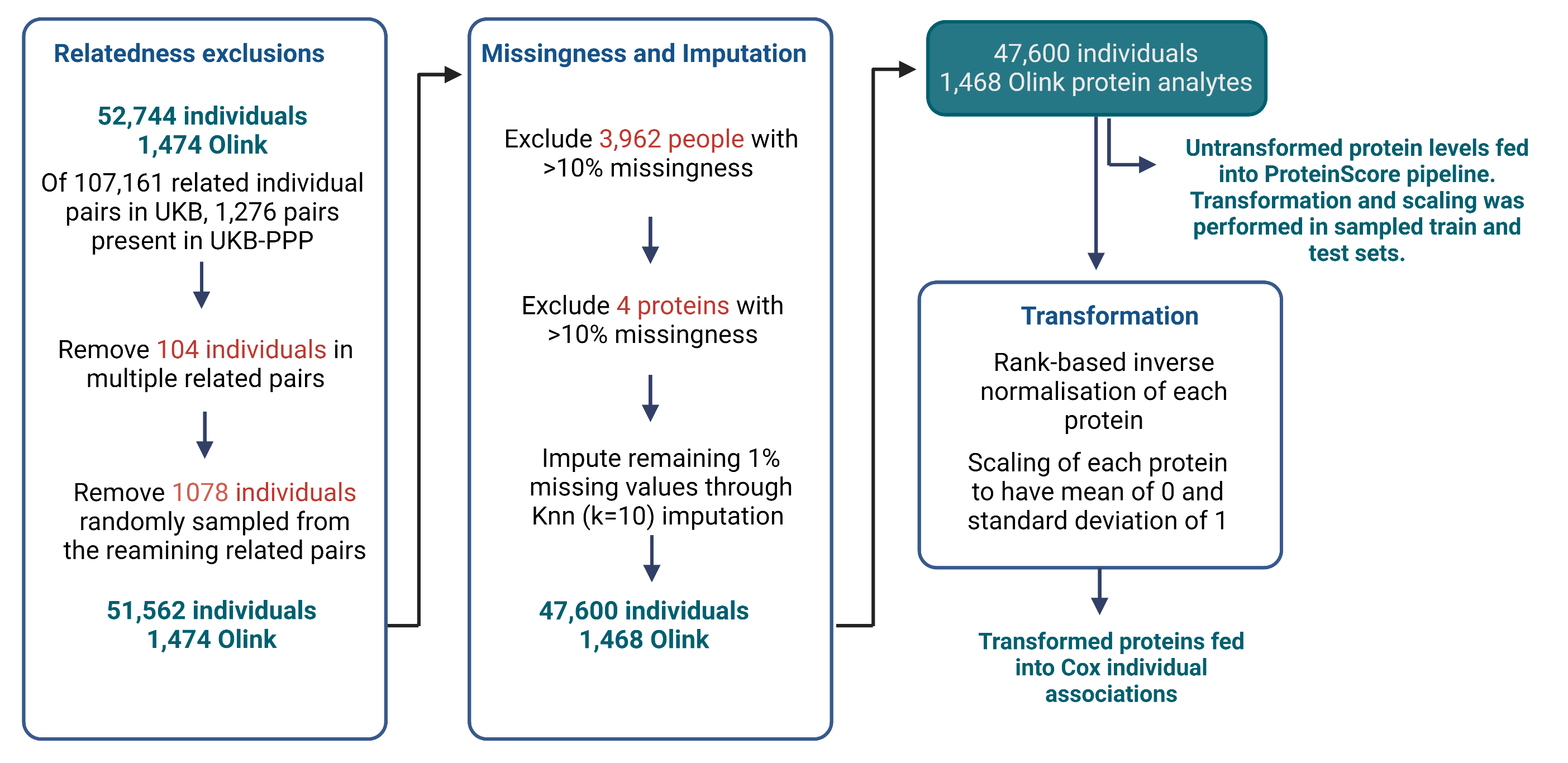

**Supplementary Figure 1. Summary of processing steps applied to the protein measurement data in UKB-PPP.** Related individuals were excluded, leaving a dataset containing 51,562 individuals with 1,474 Olink protein analytes measured. Next, 3,962 individuals that had >10% missing data were excluded, followed by four proteins that had >10% missing data. The remaining missing protein measurements (1% of total measurements) were imputed through K-nearest neighbours (Knn; k=10) imputation. The final dataset was comprised of 47,600 individuals and 1,468 Olink protein analytes. Protein levels were rank-based inverse normalised and scaled to have a mean of 0 and standard deviation of 1 prior to individual Cox PH analyses. Untransformed protein levels were fed into the model pipeline for ProteinScore development and were rank-based inverse normalised and scaled to have a mean of 0 and standard deviation of 1 in train and test sets separately once these were sampled for each outcome.

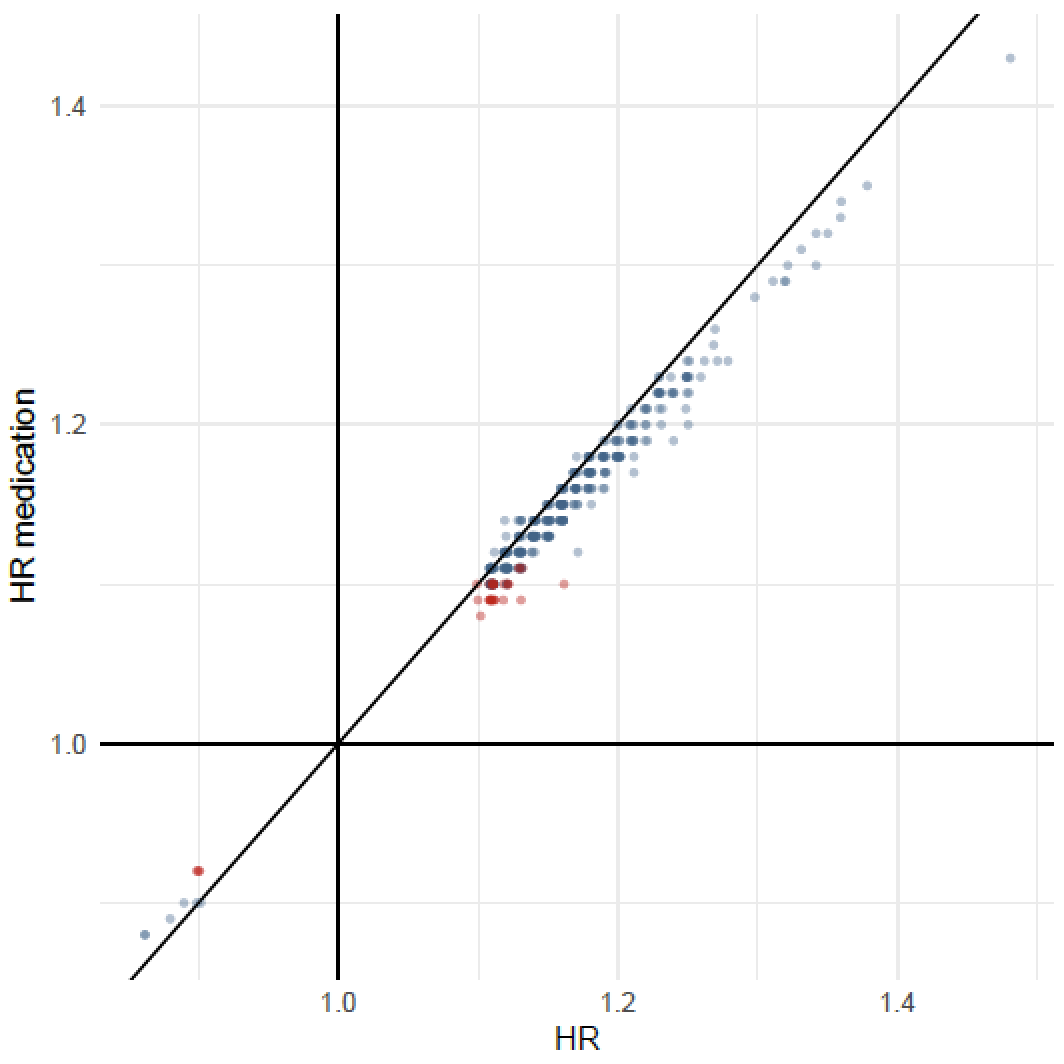

**Supplementary Figure 2. Hazard ratio replication for 370 protein associations with ischaemic heart disease in models that did (y-axis) and did not (x-axis) adjust for blood-pressure lowering medication use at baseline.** Both sets of results adjusted for age, sex and six lifestyle factors and were performed in a subpopulation of 35,073 of the 47,600 individuals that had medication information available. Of the 35,073 individuals, 14,074 reported taking one of more blood-pressure lowering medications. The 36 associations attenuated by adjusting for blood-pressure lowering medication use are highlighted in red, whereas the 344 associations that had P < 3.1x10^-6^ (Bonferroni-adjusted P-value) across both models are shown in blue.

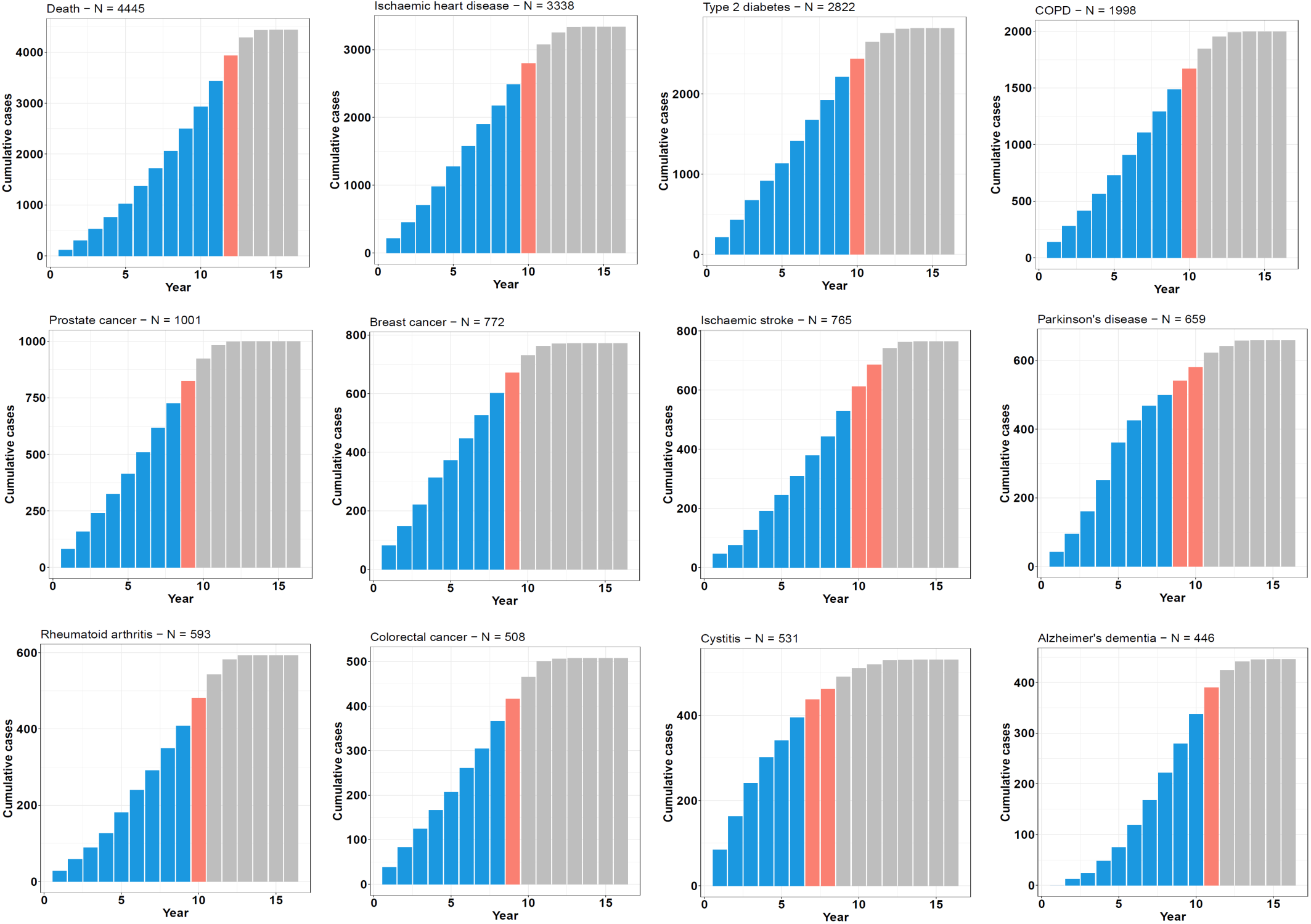

**Supplementary Figure 3. Cumulative time-to-onset for cases by outcome in the UK Biobank PPP sample.** Case counts across the 16-year follow-up period are shown for each trait, with the number of cases by year of follow-up plotted cumulatively and the year that the proportion of cases diagnosed reached 80% (orange) and 90% (grey) demarcated. COPD: chronic obstructive pulmonary disease.

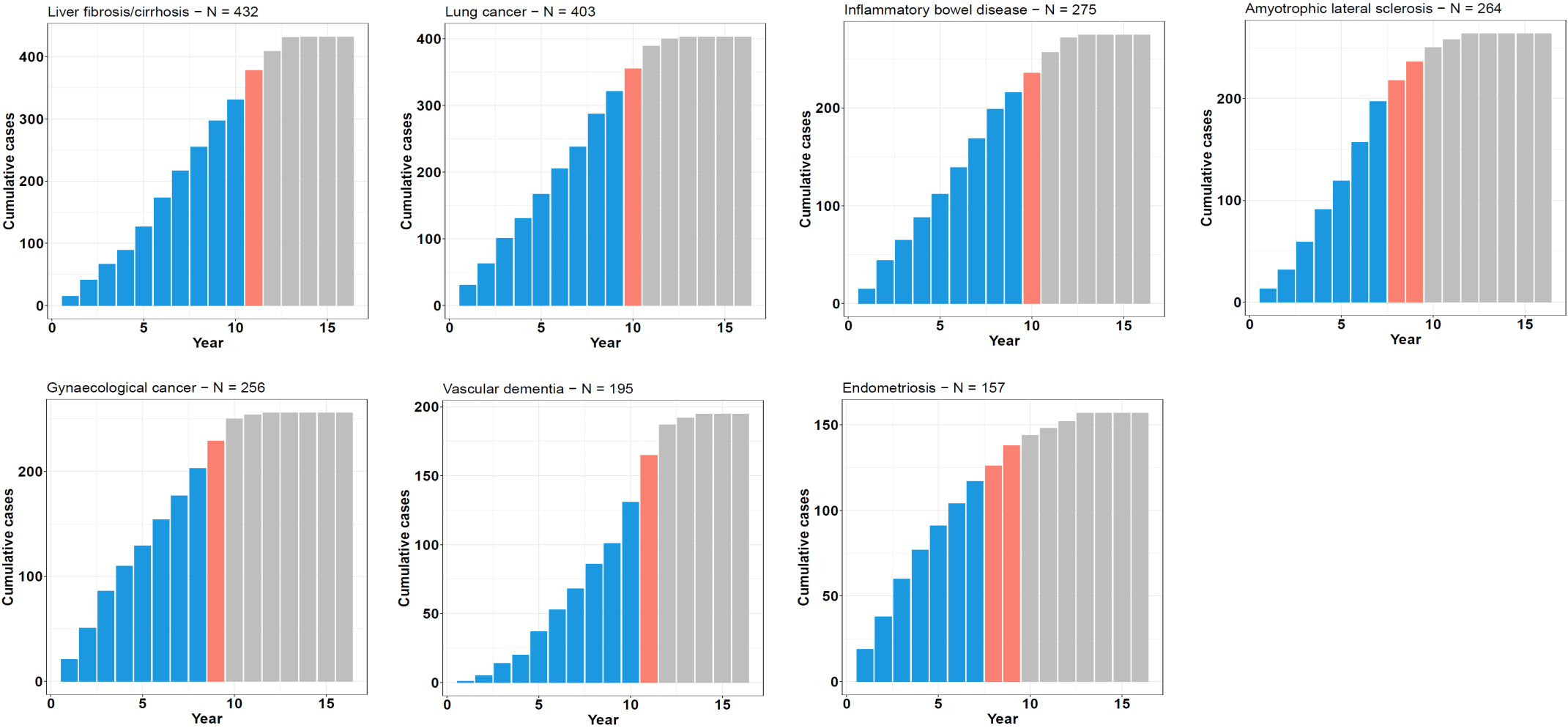

**Supplementary Figure 4. Cumulative time-to-onset for cases by outcome in the UK Biobank PPP sample.** Case counts across the 16-year follow-up period are shown for each trait, with the number of cases by year of follow-up plotted cumulatively and the year that the proportion of cases diagnosed reached 80% (orange) and 90% (grey) demarcated.

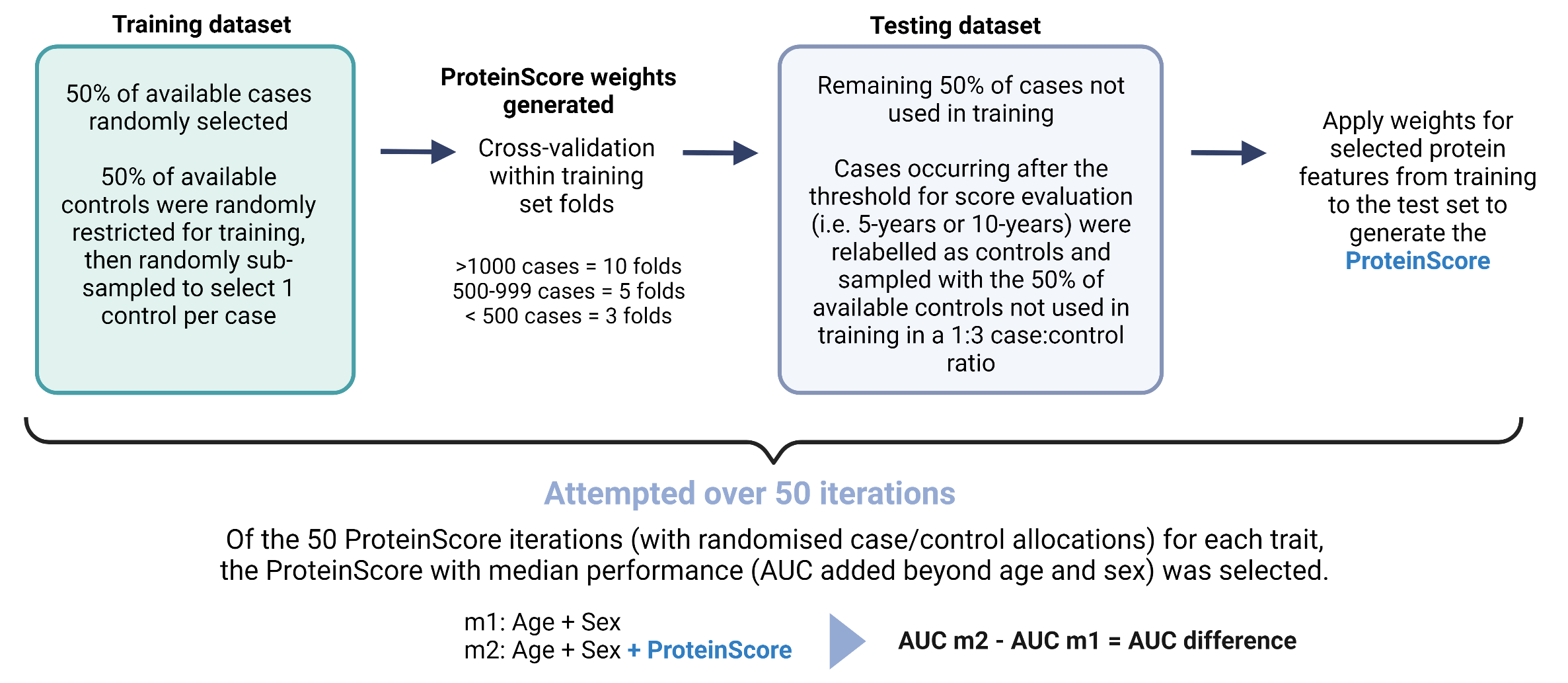

**Supplementary Figure 5. Summary of the ProteinScore development pipeline.** ProteinScores were developed across fifty randomised iterations. For each iteration, 50% of available cases were randomly allocated to the training set and 50% of controls were randomly sampled to obtain a 1:3 case:control ratio. Cox PH elastic net regression with cross-fold validation across folds of the training sample was used to derive weighting coefficients. The 50% of cases that were not included in the training set were allocated to the test set. If cases in the test set occurred after the threshold for onset evaluation (i.e. 5-year or 10-year), they were relabelled as controls and randomly sampled with the 50% of controls not considered during training, to obtain a 1:3 case:control ratio. Of the fifty ProteinScore iterations tested, the ProteinScore that yielded the median incremental difference to the Area Under the Curve (AUC) beyond a minimally-adjusted model was identified. If no features were selected for an iteration, it was weighted with a performance of 0 in median AUC selection. If features were selected for an iteration but the randomly sampled test set included no cases at or beyond the onset threshold (precluding extraction of baseline hazard at this point for AUC calculation) these models were excluded from the median ProteinScore selection.
