## Supplementary Information for "Blood protein levels predict leading incident diseases and mortality in UK Biobank"

**Sample selection**

A complete summary of the sample selection, processing and quality control details for the UK Biobank PPP proteomics samples is available in Sun *et al*, 2022 ^1^. Consortium members chose samples that were enriched for specific diseases of interest. The remainder of the population was randomly sampled through stratified selection against age, sex and recruitment centre. Day of the week of collection, deprivation index and participant ethnicity were confirmed as representative of the wider UK Biobank cohort. Full inclusion and eligibility criteria are detailed in: <https://biobank.ndph.ox.ac.uk/showcase/showcase/docs/casecontrol_covidimaging.pdf>.

**Olink protein technology**

Samples were fractioned to 850µl aliquots and stored at -80 °C. Quantification was performed at Olink Analysis Service in Sweden using Olink Proximity Extension Assay technology. Four 384-plex panels were used (cardiometabolic, neurological, inflammatory and oncology) that targeted 1,463 unique proteins. A summary of the protein analytes available with panel information is provided in **Supplementary Table 1**. A full summary of the Olink technology used to generate analyte measurements is detailed in Sun *et al*, 2022 ^1^. Panels contained dilution blocks to account for the range of proteins present. Samples were serially diluted to 1:10, 1:100 and 1:1000 and transferred to the 384-well plates, which had four blocks for each set of 96 samples. Matched antibodies are labelled with complementary oligonucleotides that bind to the target protein in the sample. Hybridization of the probes can thus be recorded through DNA amplification using polymerase chain reaction (PCR 1) to create amplicons for protein assays. Amplicons were combined across each of the four abundance groups, resulting in one well of amplicons per sample. This signal is quantified using next generation sequencing and validated using methods that have been previously reported ^2,3^.

Details of the inbuilt Olink quality control workflow for Normalized Protein eXpression (NPX) value generation can be accessed through Sun *et al*, 2022 ^1^. Briefly, the raw data generated through the Olink quality control workflow (for 54,309 individuals) underwent removal of three control, unprocessed and withdrawn samples. Filtering was then done to remove 1) measurements that had missing NPX values or other QC failures, 2) outlier values that were beyond 5 standard deviations from the mean of the first or second standardized principal component (PCs) and 3) outliers with a median NPX greater than 5 standard deviations from the mean, or an interquaritile range (IQR) greater than 5 standard deviations from the mean IQR. After these exclusions, measurements for 54,197 individuals remained with baseline samples. Finally, data points with QC or assay warnings were removed, resulting in 58,240 samples for 54,189 individuals. The baseline samples for these 54,189 individuals were taken forward for use in the present study.

**Assessment of technical and genetic effects**

To assess the potential impact of protein processing batch (0-7), study centre (1-22) and 20 genetic principal components, protein levels were regressed onto these variables and residuals were correlated with the original protein levels. Across the 1,468 proteins tested, the lowest Pearson correlation was 0.94, indicating that there was minimal influence of these factors on protein levels. Cox PH models therefore did not incorporate them as covariates. This supports previous characterisations that suggest the proteomic data in the UK Biobank PPP sample does not have pronounced plate or batch effects ^1^.

**Summary of incident disease derivation**

Cancer diagnoses were sourced from the cancer registry made available by the UK Biobank at: <https://biobank.ndph.ox.ac.uk/ukb/label.cgi?id=100092>. First occurrence traits made available by the UK Biobank were used to define non-cancer disease diagnoses, available at: <https://biobank.ndph.ox.ac.uk/ukb/label.cgi?id=1712>. First occurrence traits integrate self-report at baseline with electronic health linkage to ICD9, ICD10 and GP read2/3 codes from healthcare providers across the United Kingdom to identify the earliest date of a given diagnosis for an individual. Self-report data was recorded at the baseline clinic visit through a touchscreen and was then confirmed via verbal interview with a nurse. Any diagnoses included as ICD codes on death registry information from the UK Biobank were also integrated. The information is linked to three-digit ICD codes, which can be used to extract diagnoses for diseases (e.g. type 2 diabetes is linked to the E11 code). Underlying datasets used to derive first occurrence data can be sourced as follows:

- ICD9: <https://biobank.ndph.ox.ac.uk/ukb/field.cgi?id=41271>
- ICD10: <https://biobank.ndph.ox.ac.uk/ukb/field.cgi?id=41270>
- GP: <https://biobank.ndph.ox.ac.uk/ukb/field.cgi?id=42040>
- Death registry: <https://biobank.ctsu.ox.ac.uk/crystal/label.cgi?id=100093>
- Self-report: <https://biobank.ctsu.ox.ac.uk/crystal/label.cgi?id=100074>

The data sources used to extract diagnoses information are summarised in **Table 1**:

| **Incident outcome** | **Coding index** | **Source** |
| --- | --- | --- |
| Cystitis | N30 | First occurrence |
| Multiple Sclerosis | G35 | First occurrence |
| Brain/CNS cancer^#^ | C71,C72 | Cancer registry |
| Schizophrenia | F20 | First occurrence |
| Systemic lupus erythematosus | M32 | First occurrence |
| Endometriosis | N80 | First occurrence |
| Vascular dementia | F01 | First occurrence |
| Amyotrophic lateral sclerosis (sourced from Motor Neurone Disease)* | G12 | First occurrence |
| Inflammatory bowel disease^#^ | K51, K50 | First occurrence |
| Major depression | F33 | First occurrence |
| Gynaecological cancers^#^ | C51,C52,C53,C54,C55,C56,C57,C58 | Cancer registry |
| Alzheimer's dementia^#^ | F00, G30 | First occurrence |
| Lung cancer | C34 | Cancer registry |
| Rheumatoid arthritis^#^ | M05,M06 | First occurrence |
| Parkinson's disease | G20 | First occurrence |
| Colorectal cancer^#^ | C18,C20,C21 | Cancer registry |
| Liver disease^#^ | K70,K71,K72,K73,K74 | First occurrence |
| Ischaemic stroke | I63 | First occurrence |
| Breast cancer | C50 | Cancer registry |
| Prostate cancer | C61 | Cancer registry |
| Chronic obstructive pulmonary disease^#^ | J40,J41,J42,J43,J44 | First occurrence |
| Type 2 diabetes | E11 | First occurrence |
| Ischaemic heart disease^#^ | I20,I21,I22,I23,I24,I25 | First occurrence |
| Death | Death status (binary) | Death registry |

**Table 1. Source of UK Biobank outcomes used in the present study.** The three-digit ICD10-associated codes for cancer (sourced from the cancer registry) and disease (sourced from first occurrence mapping) traits used to identify diagnoses are provided. Death information was sourced from the death registry. * As amyotrophic lateral sclerosis (ALS) was sourced using the motor neurone disease ICD code, individuals were checked against ALS-specific codes (GP data mapped to read 3 code F1520 and ICD codes mapped to ICD10 code G12.2) to map this trait. ^#^ Duplicate entries were removed for both cancer and non-cancer disease traits, taking the earliest date of diagnosis for a given disease outcome for each individual in order to calculate time-to-event information.

**Censoring**

Censoring dates were set in accordance with the UK Biobank data providers and dates of data availability at: <https://biobank.ndph.ox.ac.uk/ukb/exinfo.cgi?src=Data_providers_and_dates>. Censoring dates were June 2022 for non-cancer diseases, December 2016 for cancer outcomes and November 2022 for death.

**Medication use**

Medication self-report at baseline was extracted using fieldID 20003 from the UK Biobank. This covers only a portion of the UKB sample (376,448 individuals), which when subset to the population of 47,600 with protein measures available in the present study results in 35,073 individuals. To model medication use, 124,198 medication name instances that were recorded in the 35,073 individuals were condensed into unified classes of action, using the anatomical therapeutic chemical (ATC) classification categories. This coding system was previously included in the GWAS of medication classes performed by Wu *et al* ^4^. The frequency of these medications grouped into 849 ATC classes in the population of 35,073 individuals is summarised in **Supplementary Table 10**. Blood-pressure lowering medication was defined using the following ATC codes:

- Antihypertensives (ATC code C02) = 803 individuals
- Diuretics (ATC code C03) = 4227 individuals
- Beta blockers (ATC code C07) = 3660 individuals
- Calcium channel blockers (ATC code C08) = 3674 individuals
- Renin-angiotensin system actors (ATC code C09) = 7288 individuals
- Statin use (ATC code C10AA) = 8351 individuals

Taken together, 14,074 individuals (of the 35,073) indicated they were taking one or more of the above blood-pressure lowering medications at baseline. This was treated as a binary variable and the comparison with/out adjustment for this variable was performed for ischaemic heart disease Cox PH associations in the subset of 35,073 individuals. Adjustments for age, sex and six lifestyle factors were included in both sets of analyses, with 2,456 cases, 27,468 controls.

**MethylPipeR R package information**

MethylPipeR is an R package that facilitates systematic and reproducible development of complex trait and incident disease predictors and is available at:
<https://github.com/marioni-group/MethylPipeR>. A user interface for the MethylPipeR package is also available at: <https://github.com/marioni-group/MethylPipeR-UI>. In previous work, we have applied MethylPipeR to incident type 2 diabetes prediction considering DNA methylation sites as informative features ^5^. However, MethylPipeR allows for Cox PH penalised regression models to be run with for any input features of interest. Input features are provided to the model in the training sample – in this case, the measurements of 1,468 protein analytes available in the UK Biobank PPP consortium sample – and the features that are predictive of the outcome are selected and assigned weighting coefficients. These coefficients can then be applied to the test sample, to project in scores and assess performance.

**ProteinScore testing covariate preparation**

Three sets of increasingly complex covariates were used to model the difference in AUC and PRAUC resulting from the addition of ProteinScores. In addition to age and sex (that were available for all individuals), an additional set of 24 covariates were considered. These included the six lifestyle covariates modelled in individual Cox PH analyses (BMI, smoking status, alcohol consumption, social deprivation, education status and physical activity). For the extended set, 18 clinically-relevant covariates were selected from the UK Biobank biomarker panel. These have previously been integrated in metabolomics prediction studies of incident disease in the UK Biobank ^6^ and represent a comprehensive set of measures that are theoretically possible to generate in clinical settings (although generation of all biomarkers is not typically done as part of clinical practice, as disease-specific biomarkers will often be tested in specific circumstances as isolated tests). When the 24 variables were considered across the 47,600 individuals, 4,163 individuals were identified that had >10% missingness and were excluded. In the remaining 43,437 individuals, none of the covariates had >10% missingness and were therefore all retained. Age, sex, six lifestyle covariates and an extended set of 18 covariates were taken forward for ProteinScore testing. Missing covariate information for continuous traits was imputed through knn imputation and these variables were log transformed, whereas categorical and binary variables were imputed through median imputation.

**Metabolomics in the UK Biobank**

Metabolomics data were available for 12,059 individuals from the population of 47,600 that had proteomic measures available. The NMR assay includes measurement of 168 metabolites and 81 ratios between different combinations of the 168 metabolites; all measures were considered as potentially-informative features for the MetaboScore. Metabolomics data were assessed using the same imputation pipeline as the protein data; nine individuals that had >10% missingness in the 249 metabolomic measures were excluded. In the remaining population of 12.050 individuals, no metabolomic measures had <10% missingness and none were therefore excluded. The dataset was imputed via knn imputation (*k*=10). Metabolomic measure names were extracted in concordance with those set by the ukbnmr R package (Version 1.5) ^7^. Metabolomic data were rank-base inverse normalised and scaled to have a mean of 0 and standard deviation of 1 in the set population subsets used for training and testing (the same approach that was taken to produce the ProteinScore in this population for comparison).
